# Racial/ethnic disparities in sleep in mothers and infants during the Covid-19 pandemic

**DOI:** 10.1101/2021.03.22.21254093

**Authors:** Maristella Lucchini, Margaret Kyle, Nicolò Pini, Ayesha Sania, Vanessa Babineau, Morgan R. Firestein, Cristina R. Fernández, Lauren C. Shuffrey, Jennifer R Barbosa, Cynthia Rodriguez, William P. Fifer, Carmela Alcántara, Catherine Monk, Dani Dumitriu

## Abstract

**Study Objectives:** To quantify the association between race/ethnicity and maternal and infant self-reported sleep health at 4 months, exploring the role of maternal depression, stress and symptoms of trauma related to the COVID-19 pandemic as potential mediators.

**Methods:** Participants were recruited as part of the COVID-19 Mother Baby Outcomes (COMBO) cohort at Columbia University (N=71 non-Hispanic White, N=14 African American (AA), N=113 Hispanic, N=40 other/declined). Data on infant sleep were collected at 4 months postpartum. A subset of 149 women also completed questionnaires assessing maternal mental health and sleep. Multivariable regressions were used to separately estimate associations of race/ethnicity and mental health with multiple sleep domains for infants and mothers adjusting for individual-level covariates.

**Results:** Compared to non-Hispanic White, Hispanic infants slept less at night (β=- 101.7±17.6, p<0.0001) and AA and Hispanic infants went to bed later (respectively β =1.9±0.6, p<0.0001, β=1.7±0.3, p<0.0001). Hispanic mothers were less likely to perceive infant sleep as a problem (β=1.0±0.3, p=0.006). Compared to non-Hispanic White mothers, Hispanic mothers reported worse maternal sleep latency (β=1.2±0.4, p=0.002), and efficiency (β=0.8±0.4, p=0.03), but better subjective sleep quality (β=-0.7±0.4, p=0.05), and less daytime dysfunction (β=-0.8±0.4, p=0.04). Maternal mental health scores were statistically significant predictors of multiple domains of maternal sleep but did not mediate the association between race/ethnicity and sleep.

**Conclusions:** Racial/ethnic disparities in maternal and infant sleep are observable at 4 months post-partum. Maternal stress, depression and symptoms of trauma related to the COVID-19 pandemic did not mediate these associations.

## INTRODUCTION

Sleep health is an essential component of general health and it is associated with numerous mental and physical health outcomes in the general population.^1,2^ The postpartum period poses unique challenges to mothers with respect to maintaining good sleep health.^3^ In the perinatal period, poor sleep has been associated with numerous maternal physical and mental outcomes, such as changes in interpersonal relationships, risk of postpartum weight retention, and perinatal mood disorders.^4–6^ For the infant, the early postnatal period is characterized by rapid development of sleep and wake patterns, marking a major neurobiological milestone. Infants’ sleep problems have been reported in up to 15–25% of infants^7^ and are a source for concerns for both parents and pediatricians. Such sleep problems have been reported to negatively affect physical, cognitive, and socioemotional development.^8–11^ In addition to the direct adverse impact on infants and young children, several studies have indicated that poor sleep in infancy and childhood also affects parental levels of stress, depression, sense of competence, and overall quality of life,^12–14^ which could further compound the negative impact on infant development.

Despite these known risks, increasing evidence indicates that we are in the middle of a sleep crisis, with 70 million American adults estimated to suffer from chronic sleep loss and sleep disorders.^15^ This crisis also applies to children such that the average child sleep duration has decreased over the last century, with children sleeping 0.75 min less per year, on average, since 1905,^16^ and almost one third of children in the United States getting a sub-optimal amount of sleep at least once a week.^17^ However, the burden of poor sleep health is not experienced equally across the U.S. population. Individuals from racial/ethnic minority backgrounds have been shown to experience worse sleep health,^18–20^ but less is known about racial/ethnic disparities in sleep in the post-partum period.^21,22^ Similar racial/ethnic disparities in sleep have also been reported for children, such that racial/ethnic minority children compared to their White counterparts are reporting to have the highest rates of insufficient sleep.^23^ A few studies have focused on the infant period and results suggest emergence of sleep disparities early on in life.^24,25^ While these studies suggest that racial/ethnic disparities in sleep exist across the life span, the potential pathways for the relationship between race/ethnicity and poor sleep have not been fully understood. One hypothesized pathway is that of psychosocial stress, which has been shown to be associated with poor sleep^26,27^ and has been reported to disproportionately affect racial/ethnic minorities.^28–30^ In addition, most recently, racial and ethnic minorities have been disproportionately affected by the COVID-19 pandemic,^31–33^ which itself represents a major psychosocial stressor.^34^

To our knowledge, no published study exists investigating racial/ethnic disparities in infant and maternal sleep during the COVID-19 pandemic. Here, we investigate racial/ethnic disparities in sleep both in infants and mothers at 4 months post-partum, leveraging data collected as part of the COVID-19 Mother-Baby Outcomes (COMBO) initiative at Columbia University Irving Medical Center. The sample of this cohort is racially/ethnically diverse and since many participants gave birth at the height of the pandemic, it represents a unique opportunity to investigate relationships among race/ethnicity, psychosocial risk factors during the COVID-19 pandemic, and mother-infant sleep.

## METHODS

### Participants and study design

Participants were recruited as part of the COMBO cohort at Columbia University Irving Medical Center (www.ps.columbia.edu/COMBO), which aims to comprehensively describe the health of mother-infant dyads during the COVID-19 pandemic. All procedures were approved by the Columbia University Institutional Review Board. Eligible mothers delivered newborns at one of two Columbia University-affiliated New York-Presbyterian Hospitals, the Morgan Stanley Children’s Hospital or the Allen Hospital in New York City (NYC). On March 22, 2020, universal SARS-CoV-2 PCR testing via nasal swab was initiated for all women admitted to labor and delivery units. Starting in July 2020, all laboring mothers were also tested for COVID-19 antibodies. All mothers who gave birth at Morgan Stanley Children’s Hospital or Allen Hospital since March 22, 2020 and who had a confirmed SARS-CoV-2 PCR positive test result during pregnancy or a SARS-CoV-2 positive antibody test result with a confirmed or suspected infection onset during pregnancy were approached for inclusion in the COVID-19 positive group of COMBO. For each enrolled COVID-19 positive mother-baby dyad, a minimum of one case-matched dyad was approached for enrollment into the control group. Control mothers must have had negative SARS-CoV-2 PCR and antibody testing at admission to labor and delivery, no history of COVID-like symptoms, and no history of a positive COVID-19 test at any point during pregnancy. Control dyads were matched based on infant sex, gestational age in two-week windows, mode of delivery, and date of birth within approximately 1 week. A subset of participants was initially approached to enroll in the prenatal arm of the COMBO study. Prior to delivery, they were asked to complete the COVID-19 Perinatal Experiences (COPE) survey to collect information on maternal mental and physical health during pregnancy. These participants were subsequently enrolled in the postnatal arm of the COMBO study after giving birth and were enrolled regardless of COVID-19 status and without case-matching. An additional group of mothers who gave birth prior to the onset of the COVID-19 pandemic in NYC, during the month of February 2020, were also approached for enrollment as a control group for pandemic-related stress during pregnancy. All surveys were administered through the Columbia University Irving Medical Center REDCap system (version 10.6.2) and participants were offered the option to complete them in English or Spanish. After delivery, mothers were invited to complete the online surveys at 1, 2, 4, 6, and 9 months of age.

### Race/ethnicity

The main exposure variable was maternal racial/ethnic background. As part of hospital intake records and through the online surveys, mothers self-reported their ethnic background as ‘non-Hispanic’, ‘Hispanic’, or ‘decline to answer’ and were also asked to specify their racial background choosing from categories recommended by the National Institutes of Health (NIH) as ‘White’, ‘Black or African American’, ‘Asian’, ‘American Indian or Alaskan Native’, ‘Native Hawaiian or other Pacific Islander’, and/or ‘Other’. We combined these 2 variables to obtain one single race/ethnicity variable as follows: non-Hispanic White (W), African American (AA), Hispanic (H), and other (O).

### Sleep and mental health measures

The primary outcomes were mothers’ report of their infant’s sleep health as measured by the Brief Infant Sleep Questionnaire (BISQ),^35^ and of their own sleep as measured by the Pittsburgh Sleep Quality Index (PSQI) questionnaire (Cronbach’s alpha 0.83).^36^

The entire cohort completed the BISQ infant sleep questionnaire. From the BISQ we extracted the following measures: 1) nighttime sleep duration, 2) daytime sleep duration, 3) total sleep duration, 4) number of night awakenings, 5) amount of wakefulness during the night, 6) sleep latency, 7) bedtime, and 8) infant’s sleep perceived as a problem by the mother.

One-hundred and forty-nine women out of the total sample of 238 women also completed the PSQI. From the PSQI we extracted the following measures: 1) subjective sleep quality, 2) sleep latency, 3) sleep duration at night, 4) sleep efficiency, 5) sleep disturbances, 6) use of sleep medication, 7) daytime dysfunction, and 8) overall sleep health. This same subset of mothers also completed the Patient Health Questionnaire-9 (PHQ-9, Cronbach’s alpha 0.851)^37^ to assess severity of depressive symptoms, the Perceived Stress Scale (PSS, Cronbach’s alpha 0.82)^38^ to assess levels of perceived stress, and the post-traumatic stress PTSD Checklist for DSM-5 (PCL-5) adapted for COVID-19 (see Supplementary Material for this adapted scale).

### Other measures

We collected information on infant sex, gestational age at birth, maternal age at delivery, and health insurance status (commercial versus Medicaid) from the electronic medical records. In addition, we defined maternal COVID-19 status, as 1) control pre-pandemic, 2) negative, 3) positive. Positive indicated that the participant had tested positive by PCR or serology at any point during pregnancy. Negative indicated that participant had not had a known COVID-19 infection at any point in pregnancy and had tested negative for SARS-CoV-2 by PCR and/or antibody testing at the time of delivery. The subset of mothers who completed the PSQI, PHQ-9, PSS, and PCL do not include any pre-pandemic control participants.

### Statistical analysis

We tested for normal distribution of continuous variables obtained from the BISQ. In case of not normally distributed variables (amount of wakefulness during the night, sleep latency, bedtime) we discretized them and transformed from continuous to ordinal. Then, we used multiple linear/ordinal/logistic regression models to assess the independent associations of maternal race/ethnicity on maternal and infant sleep variables with non-Hispanic White group as reference. Regression models were adjusted for infant sex, gestational age at birth, and age in weeks at time of assessment in Model 1. No adjustments were made for regression models predicting maternal sleep in Model 1. Given that maternal socio-economic status (SES) has the potential to be a confounder of the relationship between race/ethnicity and maternal and infant sleep, Model 2 also adjusted for health insurance status as an indicator of SES, with commercial insurance as reference group. Two additional models tested the potential effects of SARS-CoV-2 prenatal exposure versus pandemic-related stress, respectively. In Model 3, we adjusted for COVID-19 status, with control pre-pandemic mothers as reference group. In Model 4, we adjusted for delivery timing with respect to the pandemic in New York City, which had a sharp peak in March-April 2020, to understand the different impact of various phases on the pandemic on maternal and infant sleep. Time periods of the pandemic at delivery were grouped in 2 months bin based on infant date of birth: 1) pre pandemic controls, 2) March-April 2020, 3) May-June 2020, 4) July-August 2020, and 5) September-October 2020.

After Bonferroni adjustments for multiple comparison analyses, the adjusted p-values for analyses on infant and maternal sleep measures was p=0.006.

We ran successive multiple linear/ordinal/logistic regression models to assess the independent associations of PHQ-9, PSS, and PCL-5-COVID scores with maternal and infant sleep variables. We performed mediation analysis via the CMAverse R package^39^ to assess whether PHQ-9, PSS and PCL scores were mediators in the relationship between race/ethnicity and maternal and infant sleep variables. The mediation was tested only for non-Hispanic White and Hispanic mothers.

## RESULTS

Infants included in this analysis were born between 2/1/20 and 9/30/20. The racial/ethnic composition of the entire sample was 71 non-Hispanic White, 14 African American, 113 Hispanic and 40 other. Additional demographics are in Table 1. The racial/ethnic composition of the subset of mothers who also completed the PSQI and the mental health questionnaires was 44 White non-Hispanic, 8 African American, 70 Hispanic and 27 other. The subset was not significantly different from the entire subset for any socio-demographic variable, except for infant’s age at the assessment, with infants from the subset being younger (p<0.001) and COVID-19 status, since in the subset there were no pre-pandemic control mothers. Additional socio-demographics for the subset are presented in Table S1.

**Table 1.** Demographic for the overall cohort and by race

| Entire cohort | Overall | W<br>(N=71) | AA<br>(N=14) | H<br>(N=113) | O<br>(N=40) |
| --- | --- | --- | --- | --- | --- |
| **Gestational age (weeks)** | 38.68 (1.7) | 38.34 (1.9) | 38.43 (2.3) | 38.95 (1.6) | 38.83 (1.3) |
| **Age at assessment (weeks)** | 18.99 (2.40) | 19.15 (2.42) | 19.04 (2.71) | 18.79 (2.34) | 19.26 (2.48) |
| **Sex (F/M)** | 119/119 | 31/40 | 6/8 | 58/55 | 24/16 |
| **Covid status** |  |  |  |  |  |
| *Pre-pandemic Control* | 66 | 21 | 5 | 28 | 12 |
| *Negative* | 92 | 34 | 6 | 34 | 18 |
| *Positive* | 80 | 16 | 3 | 51 | 10 |
| **Maternal age (years)** | 31.65 (5.4) | 33.0 (4.6) | 33.0 (6.6) | 30.1 (5.4) | 33.1 (5.4) |
| **Medical Insurance** |  |  |  |  |  |
| *Commercial* | 130 | 64 | 8 | 30 | 28 |
| *Medicaid* | 108 | 7 | 6 | 83 | 12 |
| **Mode of delivery** |  |  |  |  |  |
| *Vaginal* | 144 | 45 | 7 | 64 | 28 |
| *Cesarean* | 94 | 26 | 7 | 49 | 12 |
| **Parity** | 1.8 (1.0) | 1.7 (1.2) | 2.1 (1.0) | 1.95 (0.9) | 1.5 (0.7) |

### Infants sleep by race/ethnicity

Table 2 displays infant sleep variables across racial/ethnic groups.

**Table 2.**
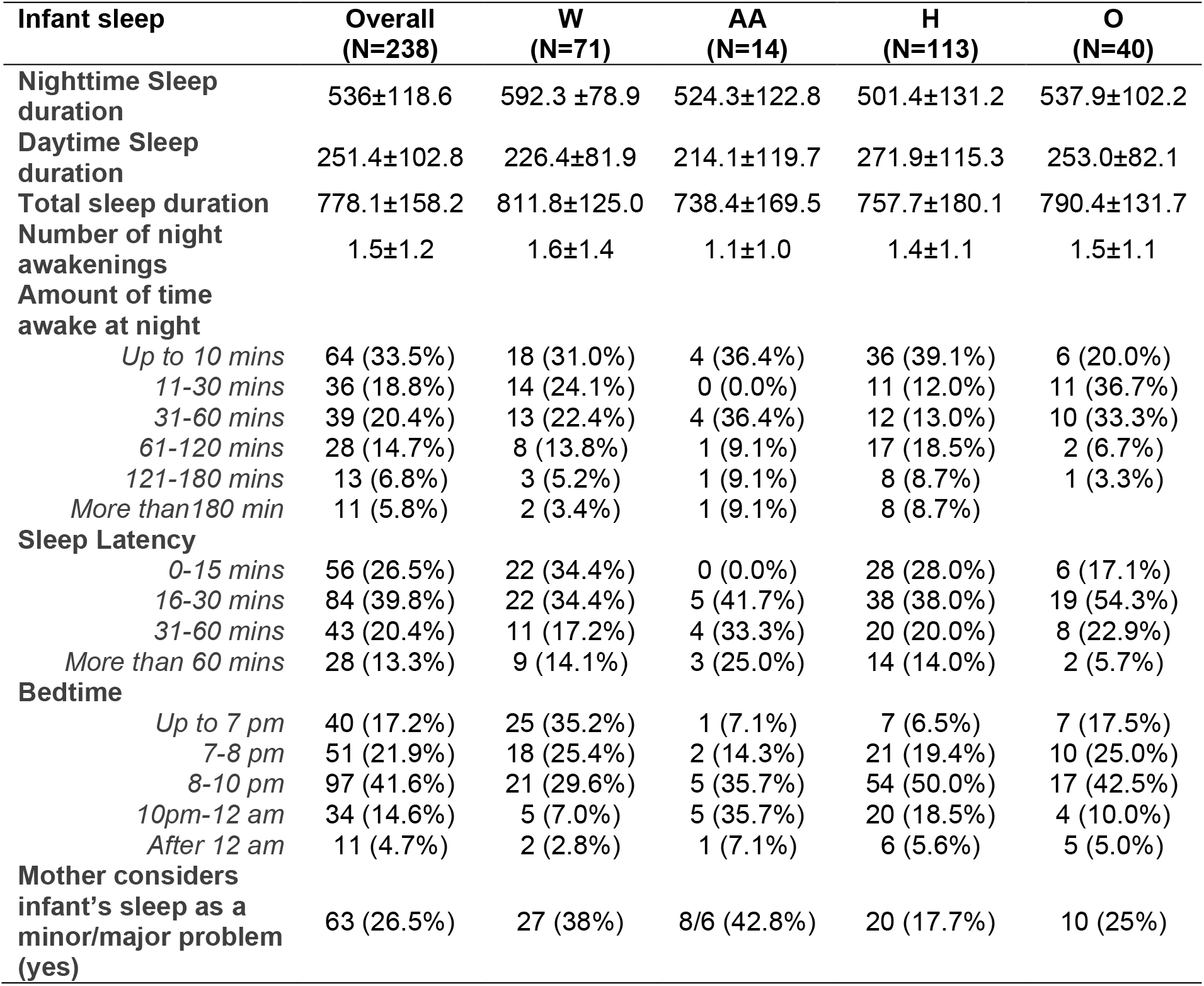
Infant sleep variables derived from the BISQ for the overall cohort and for each racial/ethnic group

The results of the multivariable regression models for Model 1, with maternal race/ethnicity predicting infant sleep variables and infant’s sex and gestational age at birth as covariates, indicated that compared to non-Hispanic White infants, Hispanic infants and infants from other race/ethnicities slept less at night, and African American infants showed the same directional trend (H: β=-101.7±17.6, p<0.0001, O: β=-64.7±23.0, p=0.005, AA: β=-63.6±33.2, p=0.06)

Compared to non-Hispanic White infants, Hispanic infants slept longer during the day (β=42.7±16.6, p=0.01), but their total sleep duration was still shorter (β=-71.5±25.2, p=0.005). Differences by race/ethnicity in nighttime, daytime, and total sleep duration are shown in Figure 1. Compared to non-Hispanic White infants, African American infants had longer sleep latency (β=1.2±0.6 p=0.03, OR 3.4 CI 1.1-10.5) and African American, Hispanic and infants from other race/ethnicities had later bedtimes (AA: β=1.9±0.6, p<0.001, OR 6.7 CI 2.2-20.2, H:β=1.7±0.3, p<0.001, OR 5.3 CI 2.9-9.8, O: β=0.9±0.4, p=0.01 OR 2.6 CI 1.2-5.3). Lastly, compared to non-Hispanic White mothers, fewer Hispanic mothers perceived their infant’s sleep as a problem (β=1.0±0.35, p=0.006 OR 2.7 CI 1.3-5.4). Figure 2 shows differences by race/ethnicity in sleep latency, bedtime, and infant’s sleep considered as a problem by the mother.

**Figure 1.**
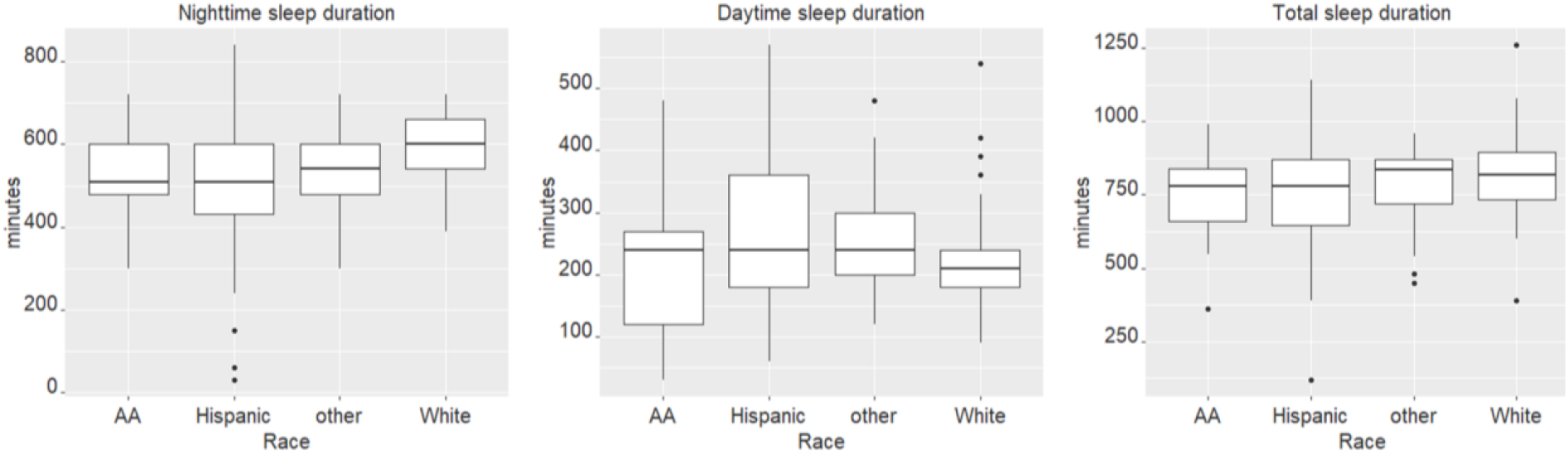
From left to right: nighttime sleep duration, daytime sleep duration and total sleep duration by race in minutes

**Figure 2.**
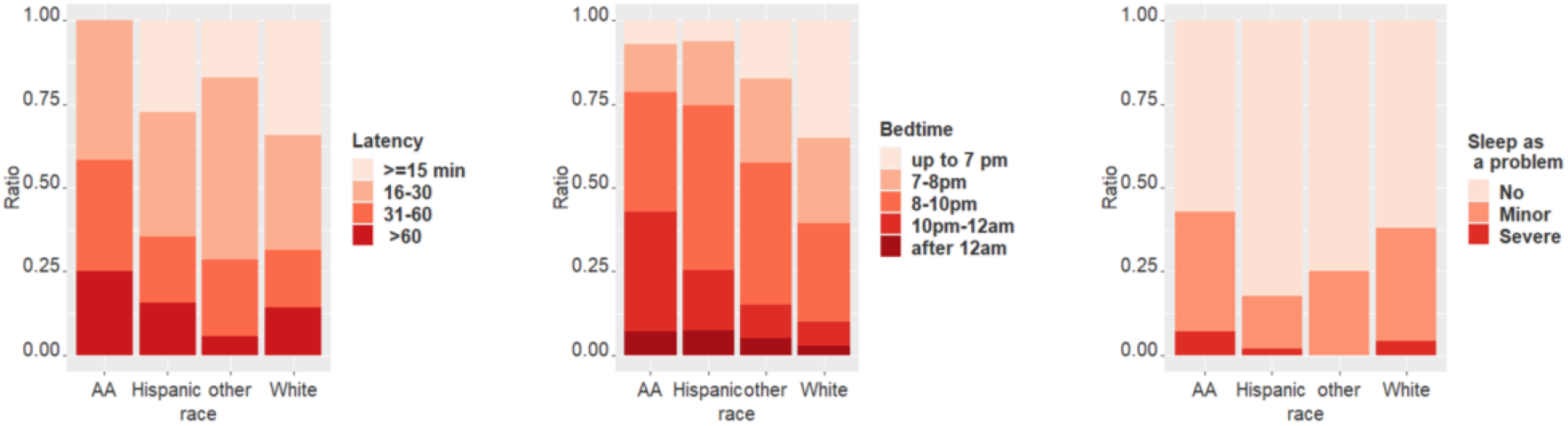
From left to right: breakdown by race of percentage of infants within each group for sleep latency, bedtime and sleep considered as a problem.

**Figure 3.**
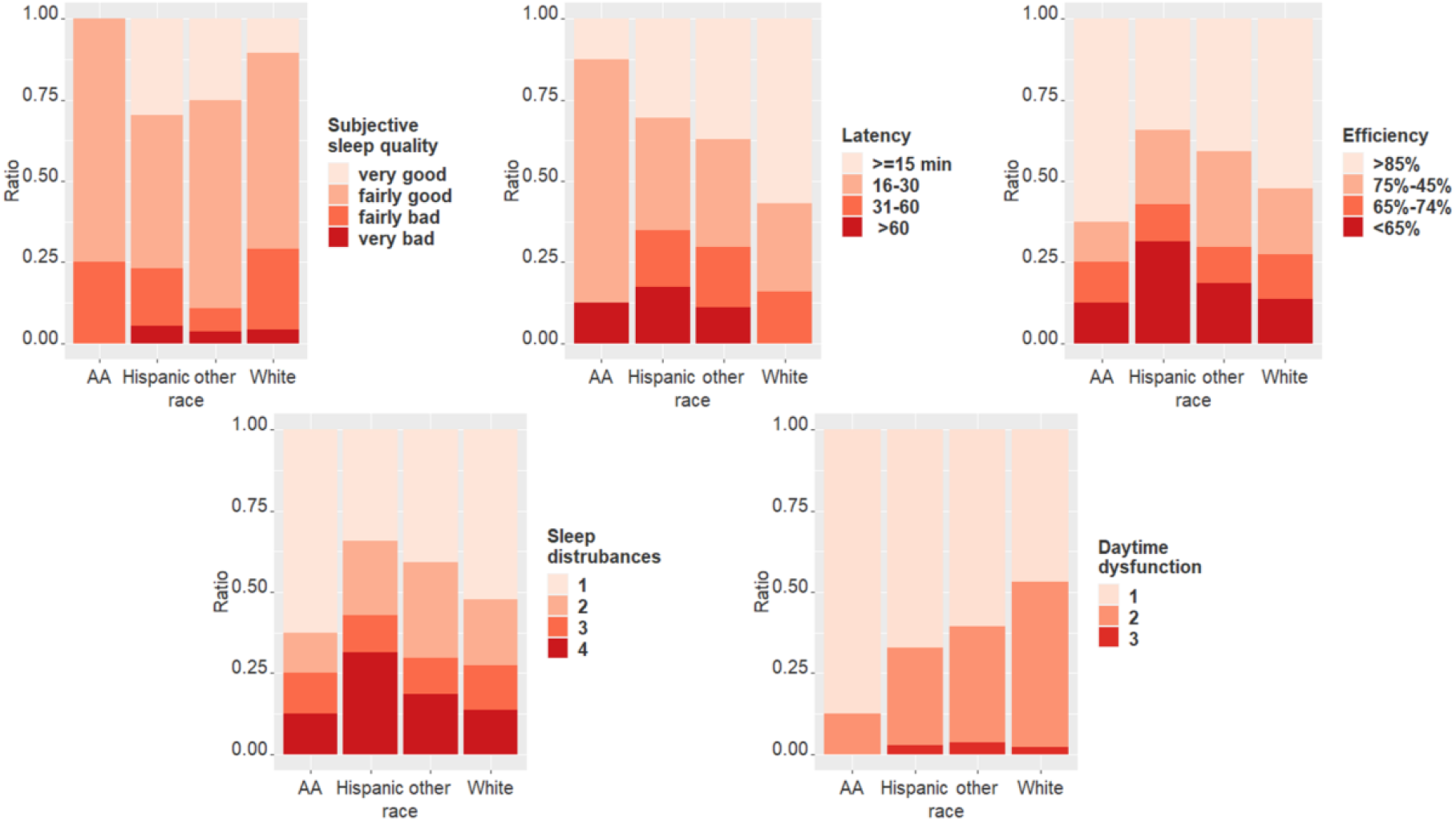
From left to right, top to bottom: breakdown by race of percentage of mothers within each group for subjective sleep quality, sleep latency, sleep efficiency, sleep disturbances and daytime dysfunction.

The covariates included in Model 1 were significant predictors for several sleep domains. Gestational age was positively associated with night time sleep duration (β=15.3±4.5, p<0.000), total sleep duration (β=14.9±6.4, p=0.02), and earlier bedtime (β=-0.25±0.07, p<0.000 OR 0.8 CI 0.7-0.9). Weeks at assessment was negatively associated with daytime sleep duration (β=- 7.3±3.0, p=0.01), number of awakenings (β=-0.2±0.06, p=0.001, odds ratio [OR] 0.8 95% confidence interval [CI] 0.7-0.9), and sleep latency (β=-0.1±0.05 p=0.02, OR 0.9 CI 0.8-1.0). Male infants woke up more during the night (β=0.6±0.3, p=0.03 OR 1.8 CI 1.1-3.0) and their mothers were less likely to report their infant’s sleep as a problem (β=-0.6±0.3, p=0.04 OR 0.5 CI 0.3-1.0).

Results from Model 2, in which we added health insurance status as an indicator of SES in the model, showed that similar differences by race/ethnicity were replicated for night and day sleep duration, latency, and bedtime. In addition, Hispanic infants had fewer nighttime awakenings compared to non-Hispanic Whites (H: β=-0.4±0.2, p=0.05). Maternal perception of infant’s sleep as a problem trended in the same direction but was no longer significant. Health insurance status was a significant predictor of bedtime, with infants from the Medicaid group going to bed later compared to infants with commercial health insurance (β=0.9±0.3, p=0.003 OR 2.4 CI 1.3-4.3).

Results from Model 3, in which we added maternal COVID-19 status, showed that the estimates associated with race/ethnicity on night and daytime sleep duration, latency, nighttime awakenings, and bedtime did not change appreciably. Maternal COVID-19 status during pregnancy was not a significant predictor of any domain of infant’s sleep at 4 months.

Results from Model 4, in which we added maternal time of delivery during the pandemic, showed that the estimates associated with race/ethnicity on night and daytime sleep duration, latency, nighttime awakenings and bedtime did not change appreciably. Time of the pandemic was associated only with sleep latency, with infants born in July-August having longer latency (β=1.3±0.6 p=0.04 OR=3.6 CI=1.0-12.9) and mothers of infants born in September-October being more likely to report infant sleep as a problem (β=-1.6±0.8 p=0.04 OR=0.2 CI=0.04-1.0). For results for Model 2, 3, and 4 see Supplementary materials for additional information on statistical results. Table 4 shows statistical results for all infant sleep domains for the 4 models.

**Table 3.** Results for regression analyses predicting infant sleep measures, for Model 1 including infant sex, gestational age at birth and age in weeks at time of assessment as covariates, Model 2 including medical insurance status as a SES indicator, Model 3 including maternal COVID-19 status and Model 4 including time of the pandemic at birth.

| Infant sleep analyses |  |  |  |  |  |
| --- | --- | --- | --- | --- | --- |
|  |  | Analyses of Variance | African American | Hispanic | Other |
| **Model 1** | Nighttime Sleep duration | F(6, 212) = 7.1, p < 0.0001 | $\beta = -63.6 \pm 33.2$, p=0.06 | $\beta = -101.7 \pm 17.6$, p<0.0001* | $\beta = -64.7 \pm 23.0$, p=0.005* |
| | Daytime Sleep duration | F(6, 205) = 2.8, p=0.01 | $\beta = -15.8 \pm 30.9$, p=0.6 | $\beta = 42.7 \pm 16.6$, p=0.01** | $\beta = 33.4 \pm 21.2$, p=0.1 |
| | Total sleep duration | F(6, 212) = 2.4, p=0.03 | $\beta = -73.1 \pm 47.6$, p=0.1 | $\beta = -71.5 \pm 25.2$, p=0.005* | $\beta = -28.2 \pm 33.0$, p=0.4 |
| | Number of night awakenings | F(6, 229) = 1.8, p=0.1 | $\beta = -0.6 \pm 0.3$, p=0.1 | $\beta = -0.2 \pm 0.2$, p=0.2 | $\beta = -0.06 \pm 0.2$, p=0.8 |
| | Amount of time awake at night | *Not applicable<sup>a</sup>* | $\beta = -0.08 \pm 0.6$, p=0.9 | $\beta = 0.2 \pm 0.3$, p=0.4 | $\beta = 0.1 \pm 0.4$, p=0.7 |
| | Sleep Latency | *Not applicable* | $\beta = 1.2 \pm 0.6$, p=0.03** | $\beta = 0.2 \pm 0.3$, p=0.6 | $\beta = 0.3 \pm 0.4$, p=0.4 |
| | Bedtime | *Not applicable* | $\beta = 1.9 \pm 0.6$, p<0.0001* | $\beta = 1.7 \pm 0.3$, p<0.0001* | $\beta = 0.9 \pm 0.4$, p=0.01** |
| | Mother considers infant's sleep as a minor/major problem | *Not applicable* | $\beta = -0.2 \pm 0.6$, p=0.7 | $\beta = 1.0 \pm 0.3$, p=0.006* | $\beta = 0.5 \pm 0.4$, p=0.2 |
| **Model 2** | Nighttime Sleep duration | F(7, 211) = 6.1, p < 0.0001 | $\beta = -62.9 \pm 33.7$, p=0.06 | $\beta = -100.2 \pm 21.2$, p<0.0001* | $\beta = -64.2 \pm 23.4$, p=0.007** |
| | Daytime Sleep duration | F(7, 204) = 2.4, p=0.02 | $\beta = -14.7 \pm 31.4$, p=0.6 | $\beta = 45.1 \pm 19.9$, p=0.02** | $\beta = 34.3 \pm 21.7$, p=0.1 |
| | Total sleep duration | F(6, 212) = 2.2, p=0.04 | $\beta = -66.8 \pm 48.3$, p=0.2 | $\beta = -57.3 \pm 30.4$, p=0.06 | $\beta = -23.5 \pm 33.5$, p=0.5 |
| | Number of night awakenings | F(7, 228) = 2.1, p=0.05 | $\beta = -0.7 \pm 0.3$, p=0.05** | $\beta = -0.4 \pm 0.2$, p=0.05** | $\beta = -0.13 \pm 0.2$, p=0.6 |
| | Amount of time awake at night | *Not applicable<sup>a</sup>* | $\beta = -0.1 \pm 0.6$, p=0.8 | $\beta = 0.03 \pm 0.4$, p=0.9 | $\beta = 0.07 \pm 0.4$, p=0.8 |
| | Sleep Latency | *Not applicable* | $\beta = 1.3 \pm 0.6$, p=0.02** | $\beta = 0.4 \pm 0.4$, p=0.3 | $\beta = 0.4 \pm 0.4$, p=0.3 |
| | Bedtime | *Not applicable* | $\beta = 1.5 \pm 0.6$, p=0.007* | $\beta = 1.2 \pm 0.3$, p=0.0007* | $\beta = 0.8 \pm 0.4$, p=0.03** |
| | Mother considers infant's sleep as a minor/major problem | *Not applicable* | $\beta = -0.3 \pm 0.6$, p=0.6 | $\beta = 0.8 \pm 0.4$, p=0.06 | $\beta = 0.4 \pm 0.4$, p=0.3 |
| **Model 3** | Nighttime Sleep duration | F(9, 209) = 4.7, p < 0.0001 | $\beta = -62.9 \pm 33.9$, p=0.06 | $\beta = -100.9 \pm 21.7$, p<0.0001* | $\beta = -64.4 \pm 23.6$, p=0.007** |
| | Daytime Sleep duration | F(9, 202) = 2.0, p=0.05 | $\beta = -13.1 \pm 31.6$, p=0.7 | $\beta = 44.2 \pm 20.2$, p=0.03** | $\beta = 33.3 \pm 21.8$, p=0.1 |
| | Total sleep duration | F(9, 209) = 1.7, p=0.08 | $\beta = -65.1 \pm 48.5$, p=0.2 | $\beta = -57.9 \pm 31.0$, p=0.06 | $\beta = -24.7 \pm 33.7$, p=0.5 |
| | Number of night awakenings | F(9, 226) = 1.6, p=0.1 | $\beta = -0.7 \pm 0.3$, p=0.05** | $\beta = -0.4 \pm 0.2$, p=0.05** | $\beta = -0.13 \pm 0.2$, p=0.6 |
| | Amount of time awake at night | *Not applicable<sup>a</sup>* | $\beta = -0.2 \pm 0.6$,<br>p=0.8 | $\beta = 0.02 \pm 0.4$,<br>p=0.9 | $\beta = 0.1 \pm 0.4$,<br>p=0.7 |
| | Sleep Latency | *Not applicable* | $\beta = 1.4 \pm 0.6$,<br>p=0.02** | $\beta = 0.4 \pm 0.4$,<br>p=0.3 | $\beta = 0.4 \pm 0.4$,<br>p=0.3 |
| | Bedtime | *Not applicable* | $\beta = 1.6 \pm 0.6$,<br>p=0.006* | $\beta = 1.1 \pm 0.3$,<br>p=0.001* | $\beta = 0.8 \pm 0.4$,<br>p=0.04** |
| | Mother considers infant's sleep as a minor/major problem | *Not applicable* | $\beta = -0.4 \pm 0.6$,<br>p=0.6 | $\beta = 0.8 \pm 0.4$,<br>p=0.08 | $\beta = 0.4 \pm 0.4$,<br>p=0.3 |
| **Model 4** | Nighttime Sleep duration | F(11, 207) = 4.1, p<0.0001 | $\beta = -62.9 \pm 34.1$,<br>p=0.07 | $\beta = -97.1 \pm 21.5$,<br>p<0.0001* | $\beta = -65.6 \pm 23.6$,<br>p=0.006* |
| | Daytime Sleep duration | F(11, 200) = 1.8, p=0.05 | $\beta = -17.6 \pm 31.8$,<br>p=0.6 | $\beta = 47.3 \pm 20.1$,<br>p=0.02** | $\beta = 30.5 \pm 21.9$,<br>p=0.2 |
| | Total sleep duration | F(11, 207) = 1.8, p=0.06 | $\beta = -72.3 \pm 48.7$,<br>p=0.1 | $\beta = -52.4 \pm 30.6$,<br>p=0.08 | $\beta = -30.0 \pm 33.7$,<br>p=0.4 |
| | Number of night awakenings | F(11, 224) = 1.4, p=0.15 | $\beta = -0.7 \pm 0.3$,<br>p=0.05** | $\beta = -0.4 \pm 0.2$,<br>p=0.04** | $\beta = -0.13 \pm 0.2$,<br>p=0.6 |
| | Amount of time awake at night | *Not applicable<sup>a</sup>* | $\beta = -0.4 \pm 0.6$,<br>p=0.5 | $\beta = 0.01 \pm 0.4$,<br>p=1.0 | $\beta = 0.06 \pm 0.4$,<br>p=0.9 |
| | Sleep Latency | *Not applicable* | $\beta = 1.2 \pm 0.6$,<br>p=0.04** | $\beta = 0.3 \pm 0.4$,<br>p=0.4 | $\beta = 0.4 \pm 0.4$,<br>p=0.3 |
| | Bedtime | *Not applicable* | $\beta = 1.6 \pm 0.6$,<br>p=0.006* | $\beta = 1.1 \pm 0.3$,<br>p=0.001* | $\beta = 0.8 \pm 0.4$,<br>p=0.04** |
| | Mother considers infant's sleep as a minor/major problem | *Not applicable* | $\beta = -0.4 \pm 0.6$,<br>p=0.5 | $\beta = 0.8 \pm 0.4$,<br>p=0.07 | $\beta = 0.4 \pm 0.5$,<br>p=0.3 |
| * p-value below 0.05 |  |  |  |  |  |
| + not significant after Bonferroni correction for multiple comparisons |  |  |  |  |  |
| <sup>a</sup> Not applicable because in ordinal regression comparisons are pairwise |  |  |  |  |  |

**Table 4.** Maternal sleep variables derived from the PSQI for the overall cohort and for each racial/ethnic group

| Maternal sleep | Overall<br>(N=149) | W<br>(N=44) | AA<br>(N=8) | H<br>(N=70) | O<br>(N=27) |
| --- | --- | --- | --- | --- | --- |
| **Subjective sleep quality** |  |  |  |  |  |
| *Very good* | 31 (20.8%) | 4 (9.1%) | 0 (0.0%) | 21 (30.0%) | 6 (22.2%) |
| *Fairly good* | 85 (57.0%) | 28 (63.6%) | 6 (75.0%) | 33 (47.1%) | 18 (66.7%) |
| *Fairly bad* | 26 (17.5%) | 10 (22.7%) | 2 (25.0%) | 12 (17.1%) | 2 (7.4%) |
| *Very bad* | 7 (4.7%) | 2 (4.6%) | 0 (0.0%) | 4 (5.8%) | 1 (3.7%) |
| **Sleep latency** |  |  |  |  |  |
| *≤15 min* | 56 (38.1%) | 25 (56.8%) | 1 (12.5%) | 20 (29.4%) | 10 (37.0%) |
| *16-30 min* | 51 (34.7%) | 12 (27.3%) | 6 (75.0%) | 24 (35.2%) | 9 (33.3%) |
| *31-60 min* | 24 (16.3%) | 7 (15.9%) | 0 (0.0%) | 12 (17.7%) | 5 (18.6%) |
| *>60 min* | 16 (10.9%) | 0 (0.0%) | 1 (12.5%) | 12 (17.7%) | 3 (11.1%) |
| **Sleep duration** |  |  |  |  |  |
| *>7* | 37 (25.5%) | 11 (25.6%) | 3 (42.9%) | 17 (24.6%) | 6 (23.1%) |
| *6-7* | 28 (19.3%) | 10 (23.3%) | 0 (0.0%) | 14 (20.3%) | 4 (15.4%) |
| *5-6* | 63 (43.5%) | 20 (46.5%) | 4 (57.1%) | 24 (34.8%) | 15 (57.7%) |
| *<5* | 17 (11.7%) | 2 (4.6%) | 0 (0.0%) | 14 (20.3%) | 1 (3.8%) |
| **Sleep efficiency** |  |  |  |  |  |
| *>85%* | 63 (42.3%) | 23 (52.3%) | 5 (62.5%) | 24 (34.3%) | 11 (40.8%) |
| *75%-84%* | 34 (22.8%) | 9 (20.5%) | 1 (12.5%) | 16 (22.9%) | 8 (29.6%) |
| *65%-74%* | 18 (12.1%) | 6 (13.6%) | 1 (12.5%) | 8 (11.4%) | 3 (11.1%) |
| *<65%* | 34 (22.8%) | 6 (13.6%) | 1 (12.5%) | 22 (31.4%) | 5 (18.5%) |
| **Sleep disturbances** | 0.8±0.5 | 0.9±0.5 | 0.9±0.6 | 0.8±0.6 | 0.7±0.5 |
| **Sleep medications** |  |  |  |  |  |
| *Not during the past month* | 137 (91.9%) | 39 (88.6%) | 6 (75.0%) | 65 (92.9%) | 27 (100.0%) |
| *Less than once a week* | 5 (3.4%) | 1 (2.3%) | 1 (12.5%) | 3 (4.2%) | 0 (0.0%) |
| *Once or twice a week* | 1 (0.7%) | 1 (2.3%) | 0 (0.0%) | 0 (0.0%) | 0 (0.0%) |
| *Three or more times week* | 6 (4.0%) | 3 (6.8%) | 1 (12.5%) | 2 (2.9%) | 0 (0.0%) |
| **Daytime dysfunction** | 0.4±0.5 | 0.5±0.5 | 0.1±0.35 | 0.3±0.5 | 0.4±0.6 |
| **Global PSQI score** | 5.9±3.4 | 5.7±3.2 | 5.6±1.8 | 6.3±3.6 | 5.5±3.4 |

### Maternal sleep by race/ethnicity

Table 3 displays maternal sleep variables across racial/ethnic groups.

The results of the multivariable regression models for Model 1, with race/ethnicity predicting maternal sleep variables indicated that compared to non-Hispanic White women, Hispanic and women from other race/ethnicities reported better subjective sleep quality (H: β=-0.7±0.4, p=0.05 OR 0.5 CI 0.24-0.99, O: β=-0.9±0.4, p=0.05 OR 0.4 CI 0.16-0.98). Nonetheless, compared to non-Hispanic White women, Hispanic women reported longer sleep latency (β=1.2±0.4, p=0.002 OR 3.2 CI 1.5-6.9), and worse efficiency (β=0.8±0.4, p=0.03 OR 2.2 CI 1.1-4.5). Women from other race/ethnicities reported less disturbances (β=-1.06 ±0.5, p=0.03 OR 0.34 CI 0.13-0.91). Hispanic women reported less daytime dysfunction (β=-0.8±0.4, p=0.04 OR 0.45 CI 0.21-0.95).

Results from Model 2, in which we added health insurance as an indicator of SES in the model, showed that the estimates associated with race/ethnicity on latency, disturbances and daytime dysfunction did not change appreciably, while for other domains differences by race/ethnicity were not significant anymore.

Results from Model 3, in which we added COVID-19 status to the model, showed that the estimates associated with race/ethnicity on latency, disturbances, and daytime dysfunction did not change appreciably. Positive versus negative COVID-19 status during pregnancy did not have any significant effects on maternal sleep measures at 4 months postpartum.

Results from Model 4, in which we added maternal time of delivery during the pandemic, showed that the estimates associated with race/ethnicity on latency and daytime dysfunction did not change appreciably. In addition, birth timing with respect to the pandemic was associated with several sleep domains. Compared to the group who delivered in March-April, the group who delivered in May-June had worse subjective sleep quality (β=1.2±0.5, p=0.02 OR 3.4 CI 1.2-10.1), longer sleep latency (β=1.3±0.6, p=0.02 OR 3.7 CI 1.3-11.5), more sleep disturbances (β=1.5±0.6, p=0.01 OR 4.4 CI 1.4-14.5), and worse overall sleep (β=1.0±0.5, p=0.04 OR 2.7 CI 1.0-7.3). Compared to the group who delivered in March-April, the group who delivered in July-August had longer sleep latency (β=1.3±0.5, p=0.01 OR 3.7 CI 1.3-10.9), more sleep disturbances (β=1.4±0.6, p=0.01 OR 3.8 CI 1.3-11.6), and worse overall sleep (β=0.9±0.5, p=0.05 OR 2.6 CI 1.0-6.5). For this analysis we did not have a control group of mothers who delivered before the pandemic.

For results for Models 2, 3, and 4 see Supplementary materials for additional information on statistical results.

Table 5 shows statistical results for all infant sleep domains for the 4 models.

**Table 5.**
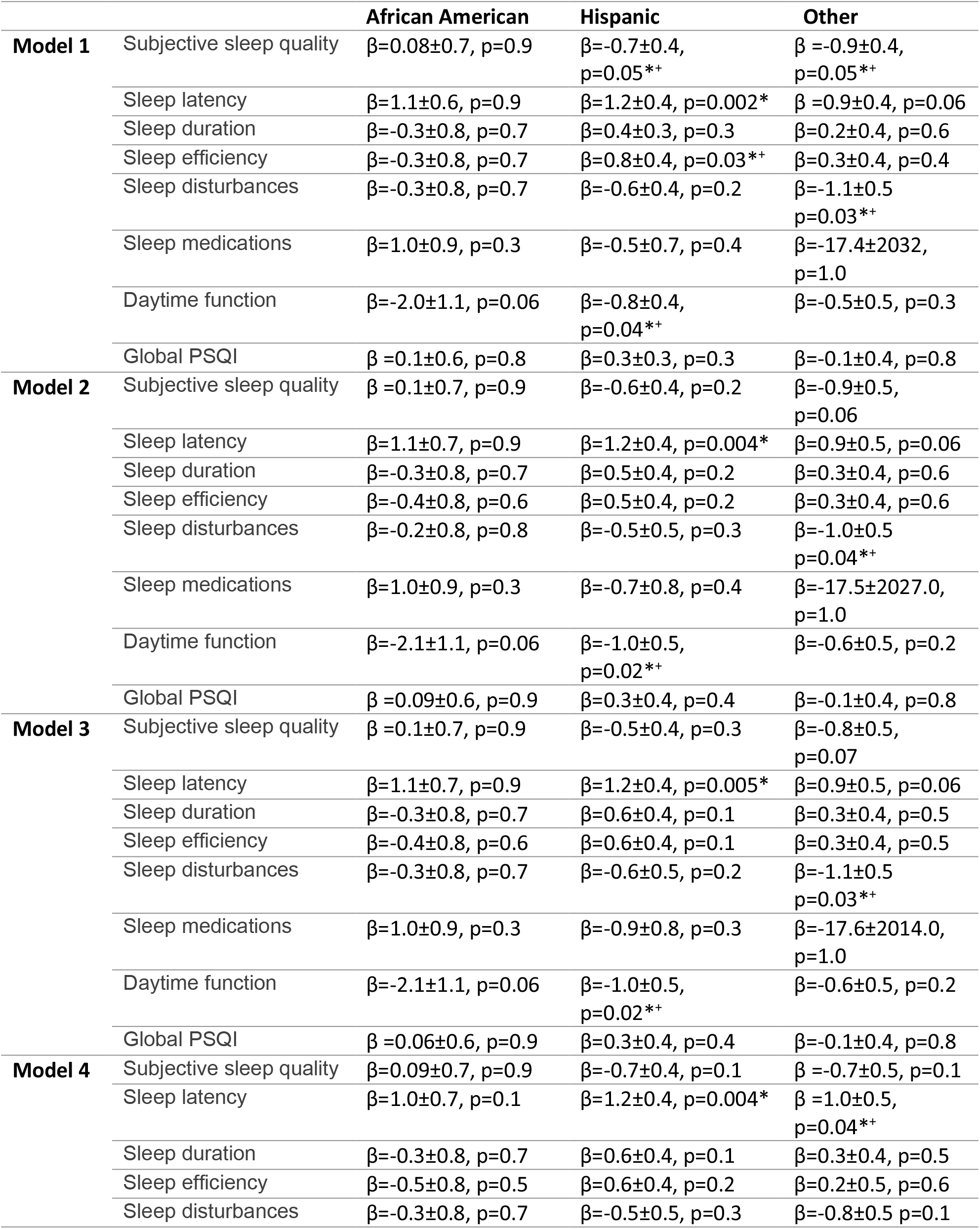
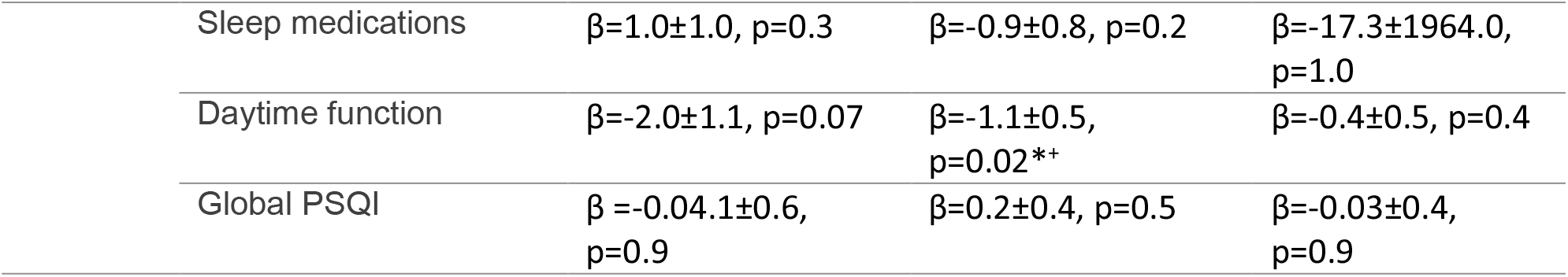
Results for regression analyses predicting maternal sleep measures, for Model 1 including no covariates, Model 2 including medical insurance status as a SES indicator, Model 3 including maternal COVID-19 status and Model 4 including time of the pandemic at birth.

|  |  | African American | Hispanic | Other |
| --- | --- | --- | --- | --- |
| **Model 1** | Subjective sleep quality | $\beta=0.08\pm0.7, p=0.9$ | $\beta=-0.7\pm0.4, p=0.05^{**}$ | $\beta=-0.9\pm0.4, p=0.05^{**}$ |
| | Sleep latency | $\beta=1.1\pm0.6, p=0.9$ | $\beta=1.2\pm0.4, p=0.002^*$ | $\beta=0.9\pm0.4, p=0.06$ |
| | Sleep duration | $\beta=-0.3\pm0.8, p=0.7$ | $\beta=0.4\pm0.3, p=0.3$ | $\beta=0.2\pm0.4, p=0.6$ |
| | Sleep efficiency | $\beta=-0.3\pm0.8, p=0.7$ | $\beta=0.8\pm0.4, p=0.03^{**}$ | $\beta=0.3\pm0.4, p=0.4$ |
| | Sleep disturbances | $\beta=-0.3\pm0.8, p=0.7$ | $\beta=-0.6\pm0.4, p=0.2$ | $\beta=-1.1\pm0.5, p=0.03^{**}$ |
| | Sleep medications | $\beta=1.0\pm0.9, p=0.3$ | $\beta=-0.5\pm0.7, p=0.4$ | $\beta=-17.4\pm2032, p=1.0$ |
| | Daytime function | $\beta=-2.0\pm1.1, p=0.06$ | $\beta=-0.8\pm0.4, p=0.04^{**}$ | $\beta=-0.5\pm0.5, p=0.3$ |
| | Global PSQI | $\beta=0.1\pm0.6, p=0.8$ | $\beta=0.3\pm0.3, p=0.3$ | $\beta=-0.1\pm0.4, p=0.8$ |
| **Model 2** | Subjective sleep quality | $\beta=0.1\pm0.7, p=0.9$ | $\beta=-0.6\pm0.4, p=0.2$ | $\beta=-0.9\pm0.5, p=0.06$ |
| | Sleep latency | $\beta=1.1\pm0.7, p=0.9$ | $\beta=1.2\pm0.4, p=0.004^*$ | $\beta=0.9\pm0.5, p=0.06$ |
| | Sleep duration | $\beta=-0.3\pm0.8, p=0.7$ | $\beta=0.5\pm0.4, p=0.2$ | $\beta=0.3\pm0.4, p=0.6$ |
| | Sleep efficiency | $\beta=-0.4\pm0.8, p=0.6$ | $\beta=0.5\pm0.4, p=0.2$ | $\beta=0.3\pm0.4, p=0.6$ |
| | Sleep disturbances | $\beta=-0.2\pm0.8, p=0.8$ | $\beta=-0.5\pm0.5, p=0.3$ | $\beta=-1.0\pm0.5, p=0.04^{**}$ |
| | Sleep medications | $\beta=1.0\pm0.9, p=0.3$ | $\beta=-0.7\pm0.8, p=0.4$ | $\beta=-17.5\pm2027.0, p=1.0$ |
| | Daytime function | $\beta=-2.1\pm1.1, p=0.06$ | $\beta=-1.0\pm0.5, p=0.02^{**}$ | $\beta=-0.6\pm0.5, p=0.2$ |
| | Global PSQI | $\beta=0.09\pm0.6, p=0.9$ | $\beta=0.3\pm0.4, p=0.4$ | $\beta=-0.1\pm0.4, p=0.8$ |
| **Model 3** | Subjective sleep quality | $\beta=0.1\pm0.7, p=0.9$ | $\beta=-0.5\pm0.4, p=0.3$ | $\beta=-0.8\pm0.5, p=0.07$ |
| | Sleep latency | $\beta=1.1\pm0.7, p=0.9$ | $\beta=1.2\pm0.4, p=0.005^*$ | $\beta=0.9\pm0.5, p=0.06$ |
| | Sleep duration | $\beta=-0.3\pm0.8, p=0.7$ | $\beta=0.6\pm0.4, p=0.1$ | $\beta=0.3\pm0.4, p=0.5$ |
| | Sleep efficiency | $\beta=-0.4\pm0.8, p=0.6$ | $\beta=0.6\pm0.4, p=0.1$ | $\beta=0.3\pm0.4, p=0.5$ |
| | Sleep disturbances | $\beta=-0.3\pm0.8, p=0.7$ | $\beta=-0.6\pm0.5, p=0.2$ | $\beta=-1.1\pm0.5, p=0.03^{**}$ |
| | Sleep medications | $\beta=1.0\pm0.9, p=0.3$ | $\beta=-0.9\pm0.8, p=0.3$ | $\beta=-17.6\pm2014.0, p=1.0$ |
| | Daytime function | $\beta=-2.1\pm1.1, p=0.06$ | $\beta=-1.0\pm0.5, p=0.02^{**}$ | $\beta=-0.6\pm0.5, p=0.2$ |
| | Global PSQI | $\beta=0.06\pm0.6, p=0.9$ | $\beta=0.3\pm0.4, p=0.4$ | $\beta=-0.1\pm0.4, p=0.8$ |
| **Model 4** | Subjective sleep quality | $\beta=0.09\pm0.7, p=0.9$ | $\beta=-0.7\pm0.4, p=0.1$ | $\beta=-0.7\pm0.5, p=0.1$ |
| | Sleep latency | $\beta=1.0\pm0.7, p=0.1$ | $\beta=1.2\pm0.4, p=0.004^*$ | $\beta=1.0\pm0.5, p=0.04^{**}$ |
| | Sleep duration | $\beta=-0.3\pm0.8, p=0.7$ | $\beta=0.6\pm0.4, p=0.1$ | $\beta=0.3\pm0.4, p=0.5$ |
| | Sleep efficiency | $\beta=-0.5\pm0.8, p=0.5$ | $\beta=0.6\pm0.4, p=0.2$ | $\beta=0.2\pm0.5, p=0.6$ |
| | Sleep disturbances | $\beta=-0.3\pm0.8, p=0.7$ | $\beta=-0.5\pm0.5, p=0.3$ | $\beta=-0.8\pm0.5, p=0.1$ |

|  |  |  |  |
| --- | --- | --- | --- |
| Sleep medications | $\beta=1.0\pm1.0$, $p=0.3$ | $\beta=-0.9\pm0.8$, $p=0.2$ | $\beta=-17.3\pm1964.0$,<br>$p=1.0$ |
| Daytime function | $\beta=-2.0\pm1.1$, $p=0.07$ | $\beta=-1.1\pm0.5$,<br>$p=0.02^{**}$ | $\beta=-0.4\pm0.5$, $p=0.4$ |
| Global PSQI | $\beta=-0.04\pm0.6$,<br>$p=0.9$ | $\beta=0.2\pm0.4$, $p=0.5$ | $\beta=-0.03\pm0.4$,<br>$p=0.9$ |

### Maternal mental health

Mothers reported an average PSS score of 18.2 (sd 7.6), with 23.8% of the mothers reporting significant levels of perceived stress (PSS score ≥25)^40^. Mothers reported an average PCL-5-COVID score of 7.7 (7.6) and an average PHQ-9 score of 2.4 (2.6), with 15.2% presenting mild levels of depression (PHQ-9 score: 5-9), 3% moderate (PHQ-9 score: 10-14) and 0.6% moderately severe (PHQ-9 score: 15-19).

Neither the PSS nor the PCL-5-COVID were significant predictors of infant sleep measures. The PHQ-9 was positively associated with maternal perception of sleep as a problem (β=0.1±0.06, p=0.05). Regarding the relationship between maternal mental health and maternal sleep, higher PSS scores were associated with poorer subjective sleep quality (β=0.08±0.02, p<0.000), longer sleep latency (β=0.06±0.02, p=0.002), more sleep disturbances (β=0.1±0.02, p<0.000), more daytime dysfunction (β=0.1±0.02, p<0.000), and overall higher global PSQI scores (β=0.08±0.02, p<0.000). Higher PCL-5-COVID scores were also associated with poorer subjective sleep quality (β=0.07±0.02, p=0.001), longer sleep latency (β=0.07±0.02, p<0.000), more sleep disturbances (β=0.1±0.02, p<0.000), more daytime dysfunction (β=0.1±0.02, p<0.000), and overall higher global PSQI scores (β=0.09±0.02, p<0.000). Lastly, higher PHQ-9 scores were associated with poorer subjective sleep quality (β=0.3±0.06, p<0.000), longer sleep latency (β=0.3±0.06, p<0.000), shorter sleep duration (β=0.2±0.06, p<0.000), more sleep disturbances (β=0.4±0.08, p<0.000), more daytime dysfunction (β=0.4±0.08, p<0.000), and overall higher global PSQI scores (β=0.4±0.06, p<0.000).

### Mediation analysis

We found no associations between race/ethnicity and scores on the PHQ-9, PSS, or PCL-5-COVID. Results from mediation analysis showed no significant result.

## DISCUSSIONS

In this prospective cohort, we found marked disparities in the sleep health of both mothers and infants by race/ethnicity at 4 months post-partum. Compared to white non-Hispanic infants, AA infants had longer sleep latency and went to bed later. Hispanic infants slept more during the day but less at night and in total and went to bed later. Observed differences in multiple sleep domains across race/ethnicity were independent of gestational age at birth, age at assessment (in weeks), and infant sex. These findings are consistent with previous literature indicating racial and ethnic differences in sleep duration in infancy.^24,41^ Extending previous results, we also report differences by race/ethnicity for several sleep domains. Later bedtime for Hispanic and AA children had been previously reported by Hake et al in preschoolers^42^, and here, we show this disparity is evident as early as 4 months of age.

Compared to non-Hispanic White women, Hispanic women reported longer latency and worse sleep efficiency. Nonetheless, they reported less daytime dysfunction and better subjective sleep quality. Previous studies on racial/ethnic disparities in sleep in the postpartum period are scarce. A study by Christian et al. looked at the effect of race and parity on sleep quality and found no effect of race postpartum.^22^ In that study, assessment occurred earlier than our study (4-12 weeks postpartum) and Hispanic women were excluded. Another study estimated normative values for postpartum sleep in a sample of majority White (93%) and married (91%) mothers from high SES and indicated that sleep duration during the first 4 months postpartum was relatively stable at approximately 7.2 h per night, and a sleep efficiency of 80% at 2 weeks postpartum increasing to 90% at 4 months.^43^ A similar study performed with a sample of African American and Hispanic women from low SES backgrounds reported shorter duration of sleep (6.5 h per night) and lower sleep efficiency of 74% at 6 weeks postpartum and 78% at 5 months.^44^ Thus, our results extend knowledge from studies of the general population which indicate that racial/ethnic-minority adults in the United States are at increased risk for poor sleep health, filling a gap in the literature regarding racial disparities in sleep during the specific post-partum period.

Although both infants and mothers from racial/ethnic minorities experience poorer sleep across multiple domains (duration, latency, efficiency, and disturbances), Hispanic mothers were less likely to describe their own sleep as poor, to report daytime dysfunction and to describe their infant’s sleep as a problem. This is in line with prior findings. A study performing a secondary analysis of the National Health and Nutrition Examination Survey 2005–2010^45^ found that although racial/ethnic minority women had poorer sleep health, they were less likely to report trouble sleeping to a physician compared to White women. Similarly, a study assessing parent perspectives of their infants’ sleep in a predominantly low SES African American sample found that while children’s average sleep duration was below recommended for their age group and overall sleep difficulty was high, most mothers reported that their children had normal sleep.^46^

The observed racial/ethnic differences in sleep are likely to be associated with several factors, that may include individual, family, home, neighborhood, cultural, and societal factors, as described by the social ecological model.^1,18,47^ In this study, we explored both SES and maternal psychosocial functioning as potential factors contributing to maternal and infant racial/ethnic disparities. SES has been suggested to contribute to children’s sleep due to quality of home environments^48^, parental education and family stress^49^, or due to parents’ work schedules. Using health insurance status as an indicator of SES, we found that low SES partially attenuated maternal disparities in sleep but did not attenuate infant disparities in sleep. A study by Patrick et al. also found that SES variables were not the primary mediators of the relationship between race and sleep in an older cohort of preschoolers.^50^ Their findings indicated that parent behaviors related to sleep, such as bedtime routine consistency, sleep associations, were the primary mediators, although they might not be completely independent from SES.

We had hypothesized that maternal depression, stress, and symptoms of trauma related to the COVID-19 pandemic would be higher in racial/ethnic minorities, thereby mediating disparities in sleep. Similarly to prior findings,^51^ all domains of maternal mental health were associated with maternal sleep domains, but not to infant sleep health. Contrary to our hypothesis, however, neither depression, stress, or COVID-19-related stress were significantly different by race/ethnicity and were not significant mediators of the sleep differences observed by race/ethnicity, when comparing non-Hispanic White and Hispanic participants.

In this study we were able to investigate maternal and infant sleep health in the post-partum period during the unique period of the COVID-19 pandemic. From our findings we conclude that maternal COVID-19 status and timing of giving birth with respect to the pandemic did not significantly influence infants’ sleep. Interestingly, however, timing of giving birth did influence maternal sleep, but in an unexpected direction: mothers who delivered after the peak of the pandemic in New York City (May to October) reported worse sleep than those that delivered at the peak of the pandemic (March to April). Thus, the acute stress related to the pandemic itself might not carry as high of a risk to sleep health as the “fatigue” experienced by this prolonged pandemic. Other studies that have investigated sleep after natural disasters like 2005 Hurricane Katrina^52^ and 2011 Great East Japan earthquake and tsunami^53^ also found long lasting effects of posttraumatic stress on sleep health. Importantly, findings from a study looking at a sample of women who were impacted by Hurricane Ike, indicated that sleep quality affected changes in perceived stress over time.^54^

Limitations of this study included the sparse information about additional behavioral and socioecological factors, such as marital status and social support, that may also interact with young children’s sleep and the absence of objective sleep measures. Another limitation is the small sample of AA mothers and infants in the cohort. Next steps for research in this topic area will include objective sleep measures for mothers and infants (e.g. actigraphy) and increasing the numbers of Black/African American mothers in the research sample.

In summary, our study shows racial/ethnic disparities in sleep in infants and mothers at 4 months postpartum across several sleep domains, and these disparities persist after controlling for SES and are not mediated by differences by race/ethnicity in maternal mental health. These results highlight important information on the existing health inequities in our sample. Given the short-term and long-term implications of poor sleep health for infant health and neurodevelopment and for maternal physical and mental health, these results underscore the need to further investigate root causes and mechanisms contributing to these inequities. This knowledge will be crucial to know where to invest resources and how to develop strategies to improve sleep in the postpartum period that account for the needs of populations experiencing health inequities.

## Data Availability

The datasets generated for this study are available on request to the corresponding author.

## ACKNOWLEDGMENTS

This study was supported by an award to DD and CM from the Columbia Population Research Center. The authors thank the participants of this study for their generous contribution to this work at the time of high uncertainty and stress.

## PREPRINT REPOSITORY

MedRxiv

## SUPPLEMENTARY MATERIAL

**Table S5.**

| Cohort with PSQI and Mental health info | Overall | W N=44 | AA N=8 | H N=70 | O N=27 | p-val |
| --- | --- | --- | --- | --- | --- | --- |
| **Gestational age (weeks)** | 38.8 (1.4) | 38.4 (1.5) | 38.2 (1.2) | 39.0 (1.5) | 38.9 (1.2) | $p=.12$ |
| **Sex (F/M)** | 74/75 | 19/25 | 3/5 | 36/34 | 16/11 | $p=.51$ |
| **COVID-19 status** | | | | | | $p=.041$ |
| Pre-pandemic Control | 0 | 0 | 0 | 0 | 0 |  |
| Negative | 84 | 30 | 6 | 31 | 17 |  |
| Positive | 65 | 14 | 2 | 39 | 10 |  |
| **Maternal age (years)** | 31.9 (5.4) | 33.7 (3.9) | 34.5 (7.8) | 30.4 (5.6) | 32.2 (5.2) | $p=.005$ |
| **Medical Insurance** | | | | | | $p>.001$ |
| Commercial | 88 | 40 | 6 | 23 | 19 |  |
| Medicaid | 61 | 4 | 2 | 47 | 8 |  |
| **Mode of delivery** | | | | | | $p=.23$ |
| Vaginal | 91 | 29 | 5 | 37 | 20 |  |
| Cesarean | 58 | 15 | 3 | 33 | 7 |  |
| **Parity** | 1.8 (1.0) | 1.8 (1.4) | 3.0 (0.0) | 1.8 (0.8) | 1.4 (0.7) | $p=.14$ |
| **PSS-14** | 18.3 (7.7) | 18.8 (8.3) | 14.6 (5.5) | 17.5 (7.8) | 20.6 (5.7) | $p=0.16$ |
| **PHQ-9** | 2.4 (2.6) | 2.7 (2.7) | 1.4 (1.5) | 2.6 (2.9) | 1.8 (1.8) | $p=0.3$ |
| **PCL5 covid adaptation** | 7.7 (7.6) | 8.2 (7.7) | 9.0 (8.1) | 7.6 (10.1) | 5.9 (5.1) | $p=0.6$ |

### Infants sleep analysis

Results from Model 2, in which we added insurance as a SES proxy in the model, showed that the estimates associated with race/ethnicity on night (AA: β=-62.9±33.7, p=0.06, H: β=- 100.2±21.2, p<0.000, O: β=-64.2±23.4, p=0.007) and day sleep duration (H: β=45.0±19.9, p=0.02), latency (AA: β=1.3±0.6, p=0.02, OR 3.8 CI 1.2-12.0), and bedtime (AA: β=1.5±0.6, p<0.007, OR 4.7 CI 1.5-14.7, H:β=1.2±0.3, p<0.000, OR 3.2 CI 1.6-6.4, O: β=0.8±0.4, p=0.03 OR 2.2 CI 1.0-4.7) did not change appreciably. In addition, African American and Hispanic infants had less nighttime awakenings (AA: β=-0.7±0.3, p=0.05, H: β=-0.4±0.2, p=0.05). Maternal perception of infant’s sleep trends same direction but not significant anymore (H:β=0.8±0.4, p=0.06, OR 2.2 CI 1-5.4). Insurance status was a significant predictor of bedtime, with infants from the Medicare group going to bed later (β=0.9±0.3, p=0.003 OR 2.4 CI 1.3-4.3).

Results from Model 3, in which we added maternal COVID-19 status, showed that the estimates associated with race/ethnicity on night (AA: β=-62.9±33.9, p=0.06, H: β=-100.9±21.7, p<0.000, O: β=-64.4±23.6, p=0.007) and day sleep duration (H: β=44.2±20.2, p=0.03), latency (AA: β=1.3±0.6, p=0.02, OR 3.9 CI 1.3-12.3), nighttime awakenings (AA: β=-0.7±0.3, p=0.05, H: β=- 0.4±0.2, p=0.05) and bedtime (AA: β=1.6±0.6, p<0.006, OR 4.9 CI 1.6-15.2, H:β=1.2±0.3, p<0.000, OR 3.2 CI 1.6-6.4, O: β=0.8±0.4, p=0.04 OR 2.2 CI 1.0-4.6) did not change appreciably. Maternal COVID-19 status was not a significant predictor of any domain of infant sleep.

Results from Model 4, in which we added birth timing with respect to the pandemic, showed that the estimates associated with race/ethnicity on night (AA: β=-62.9±34.1, p=0.06, H: β=- 97.1±21.5, p<0.000, O: β=-65.6±23.6, p=0.006) and day sleep duration (H: β=47.3±20.1, p=0.02), latency (AA: β=1.2±0.6, p=0.04, OR 3.2 CI 1.0-10.3), nighttime awakenings (AA: β=- 0.7±0.3, p=0.05, H: β=-0.4±0.2, p=0.04) and bedtime (AA: β=1.6±0.6, p<0.008, OR 4.6 CI 1.5- 14.5, H:β=1.2±0.3, p<0.000, OR 3.3 CI 1.6-6.5, O: β=0.7±0.4, p=0.05 OR 2.1 CI 1.0-4.5) did not change appreciably. Birth timing was associated only with sleep latency, with infants born in July-Aug having longer latency (β=1.3±0.6 p=0.04 OR=3.6 CI=1.0-12.9) and mothers of infants born in September-October being more likely to report infant sleep as a problem (β=-1.6±0.8 p=0.04 OR=0.2 CI=0.04-1.0).

### Maternal sleep analysis

Results from Model 2, in which we added insurance as an SES proxy, showed analogous differences by race for latency (H: β=1.2±0.4, p=0.004 OR 3.4 CI 1.5-8.1), and daytime dysfunction (AA: β=-2.1±1.1, p=0.05 OR 0.11 CI 0.006-0.7, H: β=-1.0±0.5, p=0.03 OR 0.4 CI 0.1-0.9), while for other domains differences by race were not significant anymore.

Results from Model 3, in which we added COVID-19 status, confirmed observed differences by race/ethnicity in latency (H: β=1.2±0.4, p=0.005 OR 3.1 CI 0.8-11.6), and daytime dysfunction (AA: β=-2.1±1.1, p=0.05 OR 0.11 CI 0.006-0.7, H: β=-1.0±0.5, p=0.03 OR 0.4 CI 0.1-0.9). COVID-19 status did not have any significant effects on maternal sleep measures.

Results from Model 4, in which we added birth timing with respect to the pandemic, confirmed observed differences by race/ethnicity in latency (H: β=1.2±0.4, p=0.005 OR 2.8 CI 0.7-10.6), and daytime dysfunction (H: β=-1.1±0.5, p=0.03 OR 0.3 CI 0.1-0.9). In addition, birth timing was associated with several sleep domains. Compared to the group who delivered in March-April (the height of the pandemic in New York City), the group who delivered in May-June had worse subjective sleep quality (β=1.2±0.5, p=0.02 OR 3.4 CI 1.2-10.1), longer sleep latency (β=1.3±0.6, p=0.02 OR 3.7 CI 1.3-11.5), more sleep disturbances (β=1.5±0.6, p=0.01 OR 4.4 CI 1.4-14.5) and worse overall sleep (β=1.0±0.5, p=0.04 OR 2.7 CI 1.0-7.3). Compared to the group who delivered in March-April, the group who delivered in July-August had longer sleep latency (β=1.3±0.5, p=0.01 OR 3.7 CI 1.3-10.9), more sleep disturbances (β=1.4±0.6, p=0.01 OR 3.8 CI 1.3-11.6) and worse overall sleep (β=0.9±0.5, p=0.05 OR 2.6 CI 1.0-6.5).

